# *PathCrisp*: An Innovative Molecular Diagnostic Tool for Early Detection of NDM-Resistant Infections

**DOI:** 10.1101/2024.07.09.24310126

**Authors:** Shrigouri Patil, Annes Siji, Dhrithi Mallur, Nazneen Gheewalla, Shraddha Karve, Maithili Kavathekar, Bansidhar Tarai, Mandar Naik, B. S. Kruthika, Vandana Hegde, Jayaprada Rangineni, Vaijayanti Gupta, Vijay Chandru, Bulagonda Eswarappa Pradeep, Reety Arora

**Author notes:** Corresponding author’s email addresses.

## Abstract

**Objective:** The rapid and early detection of infections and antibiotic resistance markers is a critical challenge in healthcare. Currently, most commercial diagnostic tools for analyzing antimicrobial resistance patterns of pathogens require elaborate culture-based testing. Our study aims to develop a rapid, accurate molecular detection system that can be used directly from culture, thereby introducing molecular testing in conjunction with culture tests to reduce turnaround time (TAT) and guide therapy.

**Methods:** *PathCrisp* assay, a combination of Loop-mediated Isothermal Amplification (LAMP) and CRISPR-based detection, maintained at a single temperature, was designed and tested on clinical isolates. The specificity and sensitivity of the assay was analyzed, post which the assay was compared with the Polymerase Chain Reaction (PCR) method to detect the New Delhi metallo-beta-lactamase (NDM) gene in carbapenem-resistant Enterobacteriaceae (CRE) clinical samples.

**Results:** Our *PathCrisp* assay demonstrated the ability to detect as few as 700 copies of the NDM gene from clinical isolates. Our assay demonstrated 100% concordance with the PCR-Sanger sequencing method, more commonly used. Additionally, the lack of the need for a kit-based DNA purification step, rather a crude extraction via heating, enables the direct use of culture samples.

**Conclusions:** The PathCrisp assay is precise, specific and rapid, providing results in approximately 2 hours, and operates at a constant temperature, reducing the need for complex equipment handling. In the near future, we hope that this assay can be further optimized and designed as a point-of-care test kit, facilitating its use in various healthcare settings and aiding clinicians in the choice of antibiotics for therapy.

**Plain language summary:** Resistance to Carbapenem, a last-line antibiotic for treatment, is a global threat. Timely diagnosis is critical for a clinician to decide upon the treatment. However, present available methods to detect resistance are either expensive or have longer turnaround time. Here, in this study, we aim to tackle both limitations by developing a rapid, instrument-light, point-of-care assay called *PathCrisp*. Our *PathCrisp* assay is a combination of isothermal amplification (a single temperature) and the CRISPR/Cas system used for diagnosis. This provides results within 2 hours and operates at a constant temperature. Our study validated the assay using Carbapenem-resistant Enterobacteriaceae clinical samples to detect the NDM gene, compared to the PCR and sequencing technique previously used. Furthermore, the *PathCrisp* assay can detect as few as 700 copies of target DNA when tested upon serial dilution, works on crude samples (does not require pure isolated DNA), and can detect NDM-positive samples directly from the culture.

## Introduction

Antimicrobial resistance (AMR), a silent pandemic, was associated with 4.95 million deaths in 2019^1^. The World Health Organization (WHO) reports that one in every five of these deaths occurred in children below five years, highlighting the acute demand for addressing the crisis^1^. Antibiotics like Carbapenems serve as the last treatment phase for high-risk patients infected with bacteria resistant to most β-lactamases. Carbapenems, a type of β-lactam antibiotic, possess a broad spectrum of antimicrobial activity against Gram-positive and Gram-negative bacteria and are resistant to most of β-lactamases, acting as a promising last line of antibiotics^2^. However, the absence of affordable and rapid diagnostic tools increases the risk of antibiotic overuse and contributes to the emergence of resistance ^3^. Regrettably, carbapenems experienced a similar resistance outbreak.

The emergence of carbapenem resistance represents a notable hazard to public health ^4^. WHO reports that carbapenem resistance-related mortality rates range from 26-70% globally, and the death rate was significantly higher in carbapenem resistance in comparison to susceptible infection^5^. Therefore, effective, timely diagnostics are crucial for guiding appropriate antibiotic treatment, particularly in resource-limited settings.

Carbapenemase production is currently reported to be one of the leading causes of carbapenem resistance^6^. Hence, carbapenemase genes are the best candidates for the detection of carbapenem resistance. Within carbapenemases, **N**ew **D**elhi **m**etallo-β-lactamases (NDM), a Class B metallo-β-lactamase, is known for its resistance to a wide range of β-lactam antibiotics including carbapenems^7^. Furthermore, NDM has spread globally, with India being its predicted epicentre^8^, increasing the need for early detection in the subcontinent.

Early screening of resistance patterns is crucial for treating high-risk patients. Conventional antibacterial susceptibility tests typically require 2-5 days. Commercially available tests like VITEK-2 can deliver results in a few hours but are expensive, require heavy equipment and need pure cultures, resulting in a turnaround time of 18-24 hours^9^. MALDI-TOF-based analysis can detect the pathogen and resistance within a few hours but also requires costly equipment^10^. Meanwhile, molecular tests, which are both rapid and accurate, offer an alternate detection platform. In nucleic acid-based diagnostics, thermal cycler-dependent PCR and qPCR are considered the gold standards. However, the implementation of these robust tests demands sophisticated equipment and techniques, such as qPCR and whole genome sequencing (WGS)^11^. Isothermal amplification techniques, particularly **L**oop-mediated isothermal **amp**lification (LAMP), are frequently suggested as molecular alternatives due to their elimination of the need for a thermal cycler and superior analytical sensitivity compared to gold standard PCR and qPCR methods^12^. LAMP utilizes the Bst enzyme, a strand-displacing polymerase, and 2-3 primer pairs leading to a concatemer product. However, LAMP’s low specificity often results in a high rate of false-positive outcomes^13^.

Meanwhile, CRISPR/Cas is a well-known tool in molecular biology, especially for gene editing. It primarily involves the Cas enzyme, a customizable nuclease and an engineered single guide RNA (sgRNA) with an RNA scaffold binding to the Cas enzyme and a customizable target-binding sequence^14^. Cas12a, a CRISPR/Cas variant, is renowned for activating collateral trans-cleavage of ssDNA^15^. This property can be exploited for diagnostic purposes^16^. For instance, when the Cas12a RNP complex encounters the target site, it binds and activates trans-cleavage, which can be utilized for diagnostic readouts using an ssDNA reporter with a fluorophore and quencher. The reporter emits fluorescence only when cleaved, enabling precise detection.

In this study, we introduce the *PathCrisp* assay, a combination of CRISPR/Cas12a and LAMP, specifically the *PathCrisp*-NDM variant for detecting carbapenemase NDM. This assay employs Fluorescence-Quencher ssDNA for real-time fluorescence-based detection within 2 hours. We compared it with the gold standard molecular detection method, PCR. using 49 CRE isolates sequenced with Sanger sequencing. *PathCrisp*-NDM demonstrated greater sensitivity than the PCR method and can also directly detect bacterial colonies. Overall, *PathCrisp-*NDM is a specific, sensitive, and rapid diagnostic assay that can assist in therapy at the point of need.

## Materials and Methods

### sgRNA and LAMP primers designing

NDM variant gene sequences were downloaded from databases such as the Comprehensive Antibiotic Resistance Database (CARD)^17^ and the National Center for Biotechnology Information (NCBI)^18^, and aligned using Clustal Omega^19^. The aligned sequences were then viewed using AliView^20^. sgRNA was designed in the conserved region, and six LAMP primers were designed around the sgRNA. (see Table 1 for sequences).

**Table 1:**
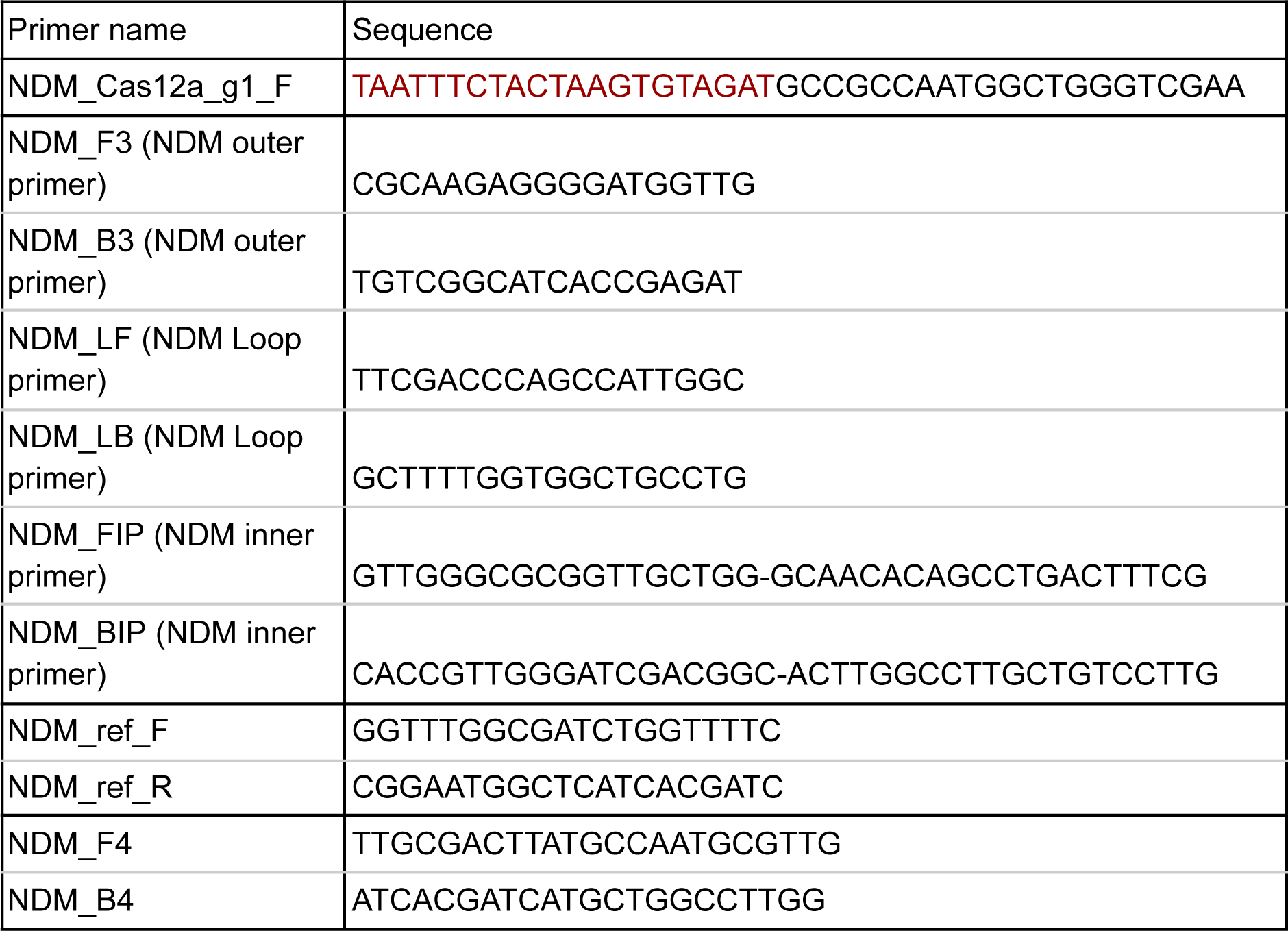
Details of the Oligo sequences used in the study.

### Testing the conservation of designed oligos and Ethical statement

This was done using the genomic sequence data from 122 isolates. Briefly, these isolates are part of a multi-centre study mapping the prevalence of AMR, as well as its genetic underpinnings, across India. With this goal, genomic DNA of three pathogenic species, namely *Escherichia coli*, *Klebsiella pneumoniae* and *Acinetobacter baumanii*, from diverse specimens were extracted in two different tertiary healthcare centers using a DNeasy Blood & Tissue Kit (69504, Qiagen).

Library was prepared using Nextera XT DNA Library Preparation Kit -96 samples (CAT No: FC-131-1096) at the sequencing facility of the National Center for Biological Sciences. Sequencing was done at the same place using the Illumina NovaSeq 6000 platform with 2×100bp sequencing read length. The raw sequence data was filtered and assembled using a GitHub pipeline^21^.

We used the fasta files of the genomic assemblies to screen for the presence of the NDM gene. For the screening, we first used ResFinder ^22^ to determine the overall antibiotic-resistance genes and their positions in the 122 assembled genomes. Using python (v 3.9.13) and RStudio (R v4.2.1) ^23^, the samples positive for NDM gene variants were screened and the NDM gene sequences (ranging from 793 to 813 base pairs in length) were extracted. Jalview (v 2.11.3.3)^24^ was used to view and align these sequences and check for the designed oligos conservation. The details of the NDM-positive isolates are given in Table S1.

Institutional ethics committee approval for the multi-centre study was obtained at Ashoka University. Additionally, both clinical partners viz. Max Healthcare and Sahyadri Hospital obtained ethical clearances from their respective ethics committees.

### Isothermal Amplification

Multi-Purpose LAMP Master Mix (DNA) (ABT-025S, Aurabiotech) was used to set up isothermal amplification. Briefly, 1 µl of the sample was added to the LAMP reaction mix containing, 5 µl of 2X reaction buffer, 0.4 µl of Bst1.0 enzyme, 0.2 µM of outer primers, 0.4 µM of loop primers, and 1.6 µM of inner primers. The reaction was incubated at 60°C for one hour.

### Trans-cleavage assay

The detection mix was prepared by pre-incubating 1.3 µM of the sgRNA, 1.3 µM of Cas12a (Alt-RTM L.b. Cas12a (Cpf1) Ultra, (10007923, IDT)), 5 µM ssDNA-FQ (/56-FAM/TT ATT

/3IABkFQ, IDT) in 1X NEBuffer™ 2 (B7002S, NEB) at room temperature for 10 mins to facilitate RNP complex formation. Following incubation, the detection mix was introduced to the template and incubated at 37^º^C for both cis and trans-cleavage activity in the Tecan Infinite MPlex with an excitation wavelength of 480 nm. Relative fluorescence units were recorded for every 2.5 minutes at 520 nm emission wavelength.

### *PathCrisp* assay-isothermal amplification followed by trans-cleavage assay (two-step)

For UDG-free amplification of the target, a Multi-Purpose LAMP Master Mix (DNA) (ABT-025S, Aurabiotech) was used. Briefly, 1 µl of the sample was added to the LAMP reaction mix containing, 5 µl of 2X reaction buffer, 0.4 µl of Bst1.0 enzyme, 0.2 µM of outer primers, 0.4 µM of loop primers and 1.6 µM of inner primers.

For amplification with UDG, WarmStart Multi-Purpose LAMP/RT-LAMP 4x Master Mix (with UDG) (M1718B, NEB) was used. Briefly, 1 µl of the sample was added to the LAMP reaction mix containing, 3.5ul of master mix, 0.2 µM of outer primers, 0.4 µM of loop primers and 1.6 µM of inner primers.

LAMP reaction was carried out at 60°C for 1 hour. No template control (NTC) was used as a negative control for the LAMP reaction in each batch.

For detection, 10 µl of amplified product was treated with the detection Mix for the trans-cleavage assay. The detection Mix was prepared as described previously in trans-cleavage assay using Cas12a (Alt-R™ A.s. Cas12a (Cpf1) Ultra, (10001272, IDT) or Alt-R^TM^ L.b. Cas12a (Cpf1) Ultra, (10007923, IDT)) and incubated at 60°C for Cis- and Trans-cleavage activity for 30 mins. An end-point reading at the 30^th^ minute was recorded in a Tecan Infinite MPlex (Sl. No: 2203009710) with 480 nm wavelength excitation and 520 nm wavelength emission.

Known NDM controls were used to study the specificity of our assay: NDM-1 positive *K.pneumoniae* (NDM-1) DNA CONTROL (MBC107-R, Vircell) and NDM negative controls: genomic DNA of *P.aeruginosa, A.baumannii, K.pneumoniae, E.coli (O157:H7)* from Himedia. Further, pJET1.2 backbone plasmid negative for NDM was used as a negative plasmid control, and a human sample’s genomic DNA was also included in the study.

No template control (Detection negative) and 0.3nM pJET1.2_NDM plasmid (Detection positive) were used in each batch as trans-cleavage controls. This pJET1.2_NDM plasmid details are mentioned in the next segment

### Limit of Detection of *PathCrisp* assay

To analyze the limit of detection of our developed assay, the NDM partial gene was amplified using NDM F4 and NDM B4 primers (sequences available in Table 1), which are located outside of outer LAMP primers, using Q5 High-Fidelity 2X Master Mix (M0492S, NEB). The amplified product was cloned into the pJET1.2/blunt plasmid using CloneJET PCR Cloning Kit (K1231, Thermo Scientific). The sequence-confirmed (sequence available in Supplementary data-1), pJET1.2_NDM plasmid was quantified using NanoDrop One™ (Thermo Scientific, USA). The concentration was 28.6 pg/µl, roughly corresponding to 7.36E+06 copies/µl (calculated through NEBioCalculator^25^). This plasmid was further 10-fold serial diluted to obtain dilutions as low as 7 copies/µl. The diluted plasmid was used to set up the *PathCrisp* assay. The assay was replicated 5 times: 3 times with Alt-R™ A.s. Cas12a (Cpf1) Ultra, (10001272, IDT) and 2 times with Alt-RTM L.b. Cas12a (Cpf1) Ultra, (10007923, IDT) at Trans-cleavage step of *PathCrisp*.

### Clinical sample preparation and Ethical statement

Clinical Samples were collected from patients, from which the pathogens were then isolated post-culture. These pathogens were further cultured, and bacterial cultures with VITEK-2 reports were obtained from Sri Venkateswara Institute of Medical Sciences, Tirupati. The DNeasy Blood & Tissue Kit (69504, Qiagen) was used for DNA extraction.

Institutional Ethics Committee approval was obtained from Sri Sathya Sai Institute of Higher Learning (SSSIHL/IEC/PSN/BS/2014/03) and Sri Venkateswara Institute of Medical Sciences Tirupati (1121 dated 20.03.2021) in accordance with the ethical standards of the Declaration of Helsinki. Further, the Institutional Biosafety Committee of C-CAMP approved the study (BT/IBKP/392/2020) and also from the MedStar Speciality Hospital Ethics Committee (Dated 07022024).

### Sequencing-based sample validation

Clinical samples were amplified using Thermo 2X PCR master mix (K0171, Thermo Scientific) as described in Mahalingam et al. 2018^26^. Briefly, a 50 µl PCR amplification was carried out using ∼20 ng template, 25 µl 2X thermo MM, and 1 µM primer mix: NDM_ref_F and NDM_ref_R. Positive samples were further sequenced to analyze the NDM variant using NCBI’s nucleotide Basic Local Alignment Search Tool (BLAST)^27^. Negative samples were further amplified by using NDM-F3/B3 and NDM-F4/B4 primers (refer to Table 1 for sequences and Table 2 for thermal cycler conditions)

**Table 2:** Details of the PCR programme used.

| Primer | Initial denaturation | Denaturation | Annealing | Extension | Number of cycles | Final extension | Expected Band size |
| --- | --- | --- | --- | --- | --- | --- | --- |
| NDM_ref_F + NDM_ref_R | 94°C for 5 mins | 94°C for 40 sec | 52°C for 40 sec | 72°C for 1 min | 30 | 72°C for 10 min | 629 bp |
| NDM_F3 + NDM_B3 | 94°C for 5 mins | 94°C for 40 sec | 53°C for 40 sec | 72°C for 1 min | 30 | 72°C for 10 min | 228 bp |
| NDM_F4 + NDM_B4 | 94°C for 5 mins | 94°C for 40 sec | 55°C for 40 sec | 72°C for 1 min | 30 | 72°C for 10 min | 334 bp |

### *PathCrisp* assay from Crude extract

To analyze the sensitivity of the *PathCrisp* assay in detecting NDM from crude extract, pJET1.2_NDM plasmid transformed *E. coli* DH5-alpha were used. Two types of culture were used: (i) streaked colony and (ii) broth culture.

For colony testing: A single bacterial colony was picked up using a 10 µl tip, dissolved in 10 µl of NFW, and boiled at 95°C for 5 mins to prepare the crude extract. One microlitre of crude extract was used to initiate *PathCrisp* assay-based diagnosis. For the media control, the pipette tip was scratched on a fresh agar plate to represent no colony control. The plasmid transformed E. coli DH5-alpha negative for pJET1.2_NDM was used for the negative control.

For broth testing: two hundred microlitres of overnight cultured broth were spun down at 13000 g for 5 minutes. Ten microliters of the culture from the bottom of the tube were collected in a fresh tube and boiled at 95°C for 5 minutes to prepare crude extract. One microlitre of crude extract was used to initiate *PathCrisp* assay-based diagnosis. For the broth-only control, 10ul of fresh LB broth was used. For the negative control, Salmonella culture was used. All the work was done in appropriate biosafety cabinets with appropriate controls.

### Diagnostic Testing Accuracy of *PathCrisp* Assay and PCR

Sensitivity and Specificity were calculated according to Shreffler J, Huecker MR, 2024 ^28^. Briefly, Sequenced confirmed samples were considered True positives and the rest were True negatives. Based on this, the following counts were made for each type of test: True positives (A), False positives (B), False negatives (C) and True negatives (D).

Sensitivity (in percentage) was calculated using the following equation:

Sensitivity= 100*(A)/((A)+(C))

Specificity (in percentage) was calculated using the following equation:

Specificity= 100*(D)/((D)+(B))

### Analysis

To determine if the sample is positive or negative, its RFU value was compared with the LAMP_NTC. In the case of multiple repeats, the Sum of 3 times the Standard deviation and average RFU value of LAMP was considered to mark the cutoff.

## Results

### Conserved regions used to design sgRNA and LAMP primers

Sequences of NDM variants 1 to 43, except for NDM-32, were analyzed for conserved regions (Figure 1.a). Details of the sequences used are available in Table S.1. The sequence for NDM-32 could not be found in any gene repository, as previously reported and hence not used ^29^. The Cas12a guide was designed to target a region conserved across these variants (Figure 1.a). LAMP primers were also built around this guide target using NDM-1 as the primary reference (Figure 1.c). SNPs that may potentially overlap the designed LAMP primers were analyzed (Figure 1.a). Outer primers: LAMP_F3 had the most SNPs and it was 51% conserved, while LAMP_B3 was 93% conserved. Inner primers: FIP (complementary of LAMP_F1 - LAMP_F2) was 97% conserved, while BIP (complementary of LAMP_B1 - LAMP_B2) was also 97% conserved. Loop primers: LAMP_LF, were also 97% conserved. LAMP_LB was 100% conserved.

**Figure 1:**
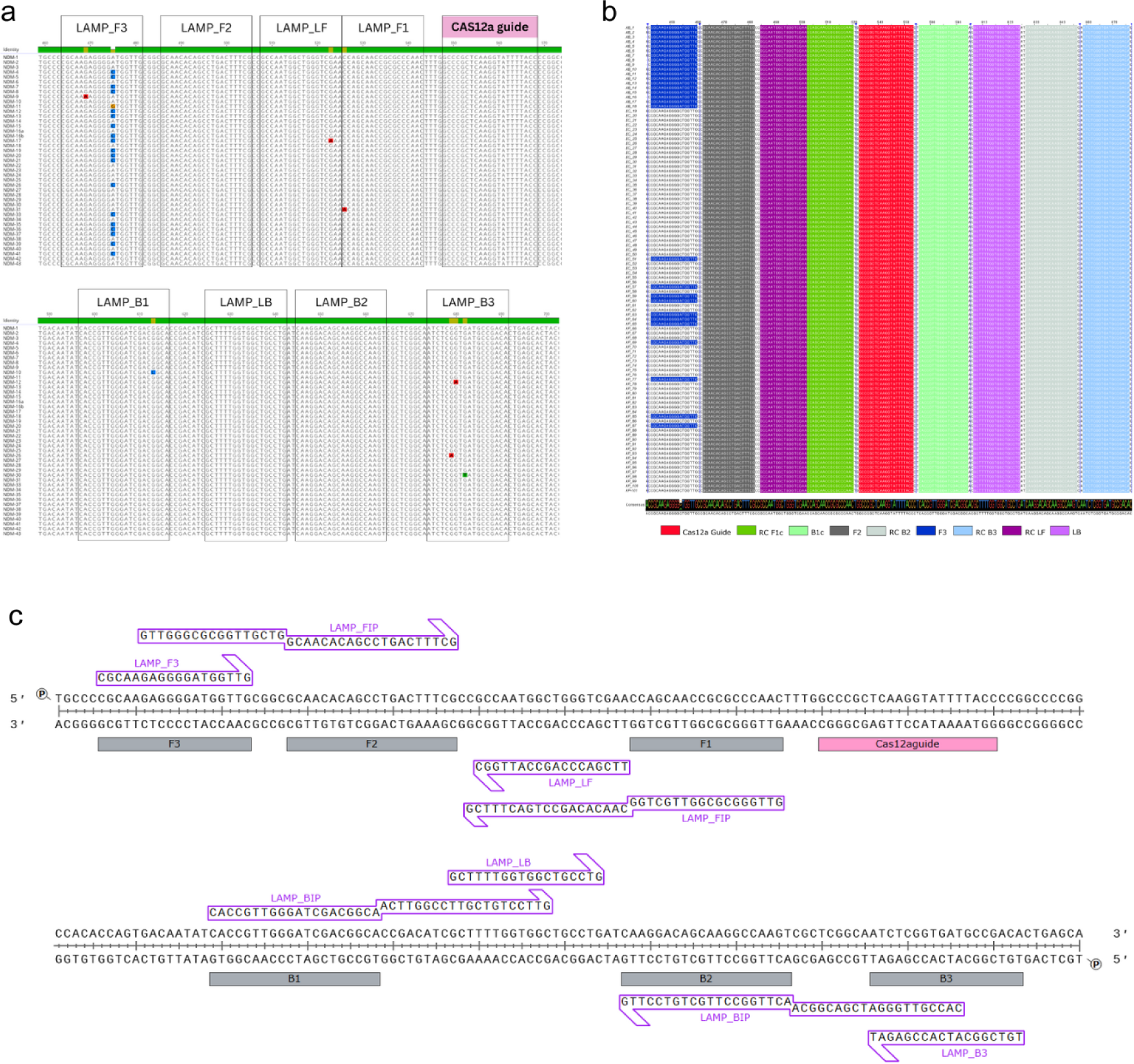
Similarity of NDM-1 variant with different variants: Primer and guide design. **a. Conserved regions across deposited NDM variants:** Partial NDM gene sequences from NCBI and CARD were aligned to study conserved regions across NDM variants (NDM-1 to NDM-43, except NDM-32). The green-coloured histogram at the beginning of the alignment depicts the conserved region. The Cas12a guide target is marked in pink, and is conserved across all variants. SNPs in the sequences are marked with block colours: dATP(red), dGTP(yellow), dCTP(blue), dTTP (green). **b. Conservation analysis in Indian patient samples:** The NDM gene sequence of 101 NDM-positive clinical samples from different priority pathogens, such as *E. coli, K. pneumoniae,* and *A. baumannii*, from Indian patients, were screened to check for the conservation of the designed oligos. Whole genome sequences of these samples were used to extract the NDM gene sequences (793–813 base pairs in length). The NDM gene regions were screened to detect homology with the designed Cas12a guide and LAMP PCR primer sequences. Abbreviations: EC - E. coli, KP - K. pneumoniae, AB - A. baumannii, RC - reverse complementary sequence. **c. Map of LAMP primers designed with Cas12a guide:** The layout of the designed LAMP primers with the sgRNA.

To further validate the conservation of these designed oligos, we screened the genomic sequence data of 122 isolates from two tertiary healthcare centers in India. 101 isolates showed the presence of NDM gene out of the total isolates screened (Fig1.b). 36 of *E.coli* out of 44, 47 of *K.pneumoniae* out of 64 and 18 *A.baumanii* out of 29 showed at least one NDM variant. All the isolates showed resistance to at least one carbapenem; while 22 isolates were resistant to all four carbapenems. Apart from LAMP_F3, all other regions were conserved. The F3 primer is conserved only in the NDM-1 variants (28.7% of the samples). The NDM-5 and NDM-4 variants (71.3% of the samples) contain an A to C SNP in this region and thus do not show homology with the F3 primer. Details of the sequences used in the study are available in Table S.1.

### Preliminary testing of designed oligos confirms their accuracy for NDM variants

To validate the designed guide and LAMP primers, a known control (genomic DNA of *K.pneumoniae* positive for NDM-1) was used to set up a 2-step assay, LAMP at 60^º^C followed by Transclevage assay at 37^º^C (Fig 2.a). The designed guide and primers were able to confirm the NDM-positive test sample. Further, to test the accuracy of our assay, different negative samples were tested in duplicates, using previously used *K.pneumoniae* (NDM-1) as assay controls (Fig 2.b). In both cases, only positive samples had higher fluorescence readings, more than 20 times of negative control (LAMP_NTC).

**Figure 2:**
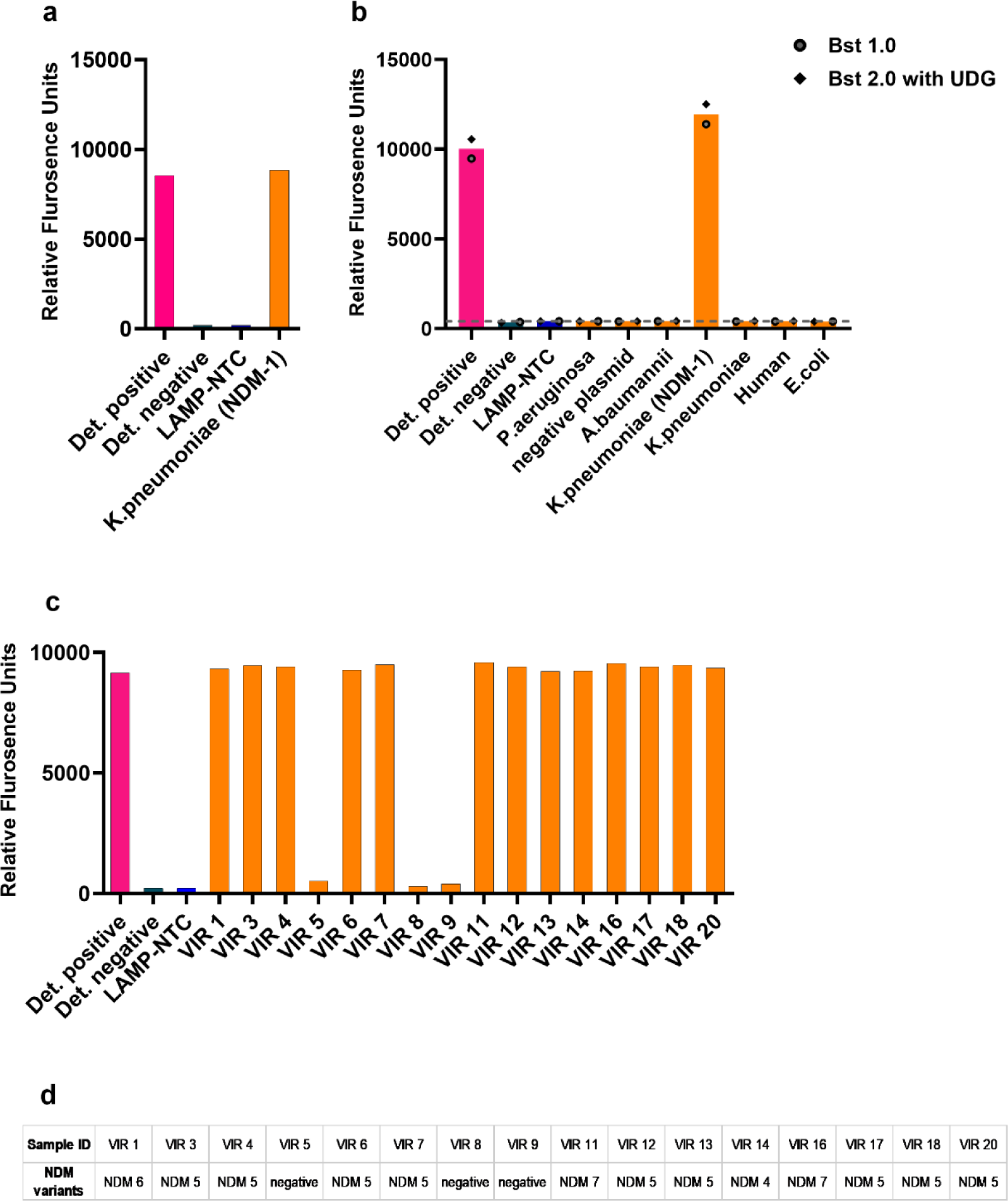
Validation of designed oligos for NDM variants: a. Testing with Genomic DNA of *K.pneumoniae:* The genomic DNA of *K.pneumoniae*, positive for NDM-1 (obtained from Vircell), is being used to test the designed LAMP primers and sgRNA. Our designed guide and primers can detect NDM-1. b. Specificity Testing with negative controls: Different known negative genomic DNAs are being used to test the specificity of the *PathCrisp* assay, with previously used *K.pneumoniae* (NDM-1) as assay control. The assay was repeated twice: once with Bst1.0 (without UDG) depicted as circles in the graph, and a second time with BST2.0 (with UDG) depicted as diamonds in the graph. The average value of each sample is represented as a bar graph, while individual values are marked either as circles or diamonds. Grey dotted lines represent the cutoff to consider if the sample is positive, which is the average of RFU of LAMP-NTC plus 3 times the standard deviation. c. Testing with Carbapenem-resistant *E.coli* Isolates: Sixteen carbapenem-resistant *E.coli* isolates from patient samples were tested with designed LAMP primers and sgRNA. VIR-5, VIR-8 and VIR-9 were negative for NDM, consistent with the previous report. d. Sequencing details of these 16 samples: Sequencing details of these 16 samples as reported previously. In each assay, the negative control for LAMP is no-template control, depicted as LAMP-NTC. For detection controls, negative control, with no LAMP product, is shown as Det. negative in the graph. For positive control, pJET1.2_NDM was used, depicted as Det. positive.

Previously, Mahalingam, Niranjana et al. (2018) had analysed AMR genes in carbapenem-resistant *E. coli*^26^. We used sixteen samples from this report to test the ability of the designed primers and sgRNA to detect different NDM-variants (Fig 2.c) and compared them with the previous data (Fig 2.d). Among 16 VIR samples, 13 were positive for NDM variants ranging from NDM-4 to NDM-7 and 3 were negative for NDM. Our assay accurately reproduced the same results as previously published, correctly calling out the NDM positives. As previously described, many NDM variants had SNPs in the LAMP F3 primer region like NDM-4 and NDM-5, yet our assay was able to specifically detect them.

### Limit of detection of *PathCrisp* assay

Our *PathCrisp* assay, integrating LAMP and CRISPR/Cas12a-based trans-cleavage, was developed to operate at a constant temperature of 60°C. This assay was meticulously analyzed to establish its limit of detection. To achieve this, a serial dilution of the pJET1.2_NDM plasmid was employed. The assay demonstrated high sensitivity, consistently detecting as little as 2.86E-03 picograms of the plasmid DNA, approximately equivalent to 700 copies of the target gene. This low detection limit highlights the assay’s potential for rapid and accurate molecular diagnostics, making it a robust tool for identifying antimicrobial resistance markers (Fig 3).

**Figure 3:**
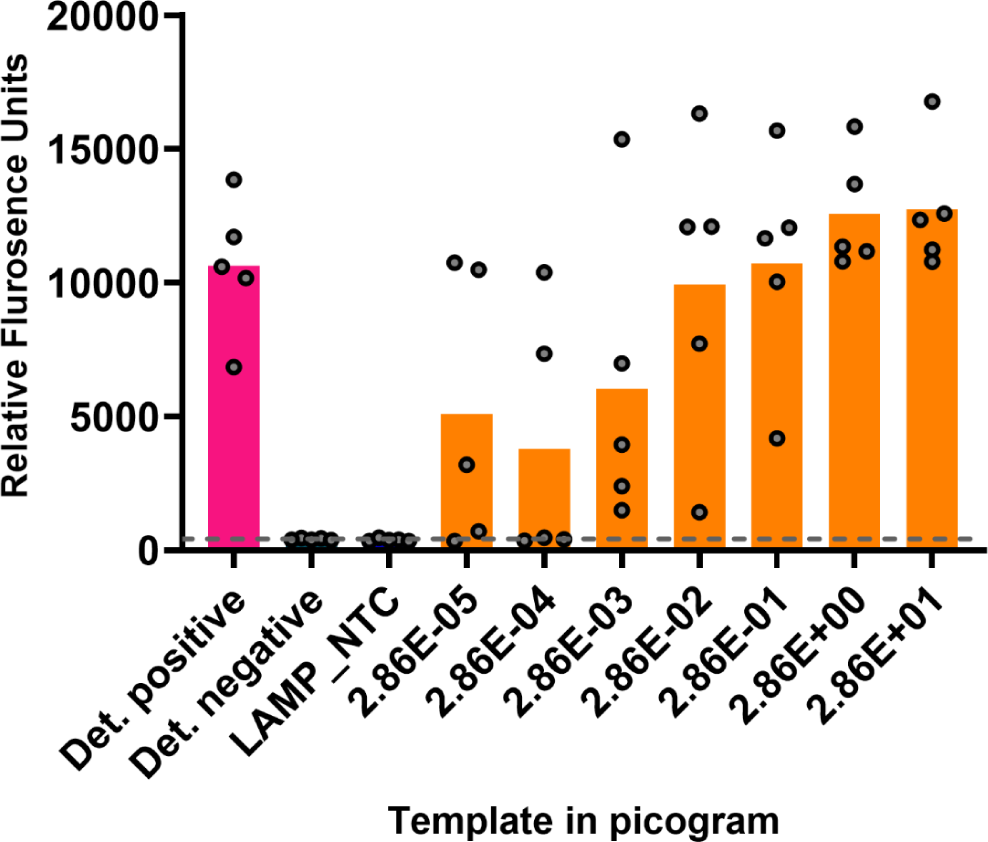
Limit of detection of NDM*-PathCrisp* assay. Purified pJET1.2_NDM plasmid was serially dilated up to 2.86E-05 picogram (which roughly corresponds to 7 copies) and then used as a template for *Pathcrisp*. The assay can consistently detect up to 2.86E-03 picogram (∼700 copies). The mean value for each sample is represented as a bar graph while individual values are marked either as circles The Grey dotted line represents the cutoff to consider if the sample is positive, which is the sum of the average LAMP-NTC RFU and 3 times its Standard deviation. The negative control for LAMP is no-template control, depicted as LAMP-NTC. For detection controls, negative control, with no LAMP product, is shown as Det. negative in the graph. For positive control, pJET1.2_NDM was used, depicted as Det. positive.

### Validation of *PathCrisp* assay using unknown CRE samples confirms its precision

Fifty Carbapenem-resistant Enterobacteriaceae (CRE) isolates were collected from patients at Sri Venkateswara Institute of Medical Sciences, Tirupati. Details about samples with VITEK data are available in Table S2. Sample BF-1302 could not be cultured and was therefore excluded from further analysis. We tested 49 samples, using the *PathCrisp* assay in batches of seven samples each, as shown in Fig 4. Among 49 samples, only sample CRE-27 was negative for NDM. To validate the assay, samples were also tested using the PCR method with electrophoresis-based detection (Figure S1). Positive bands were given for sequencing to confirm the results. However, few samples had non-specific bands or no band: samples CRE-22, CRE-23, CRE-27, CRE-33, CRE-36, CRE-47, CRE-48. To confirm the presence of the NDM gene, discrepant samples were amplified using three different sets of primers (Fig S2) and confirmed by Sanger sequencing (Table S3). Sequencing results confirmed that, except for CRE-27 all other samples were positive for NDM.

**Figure 4:**
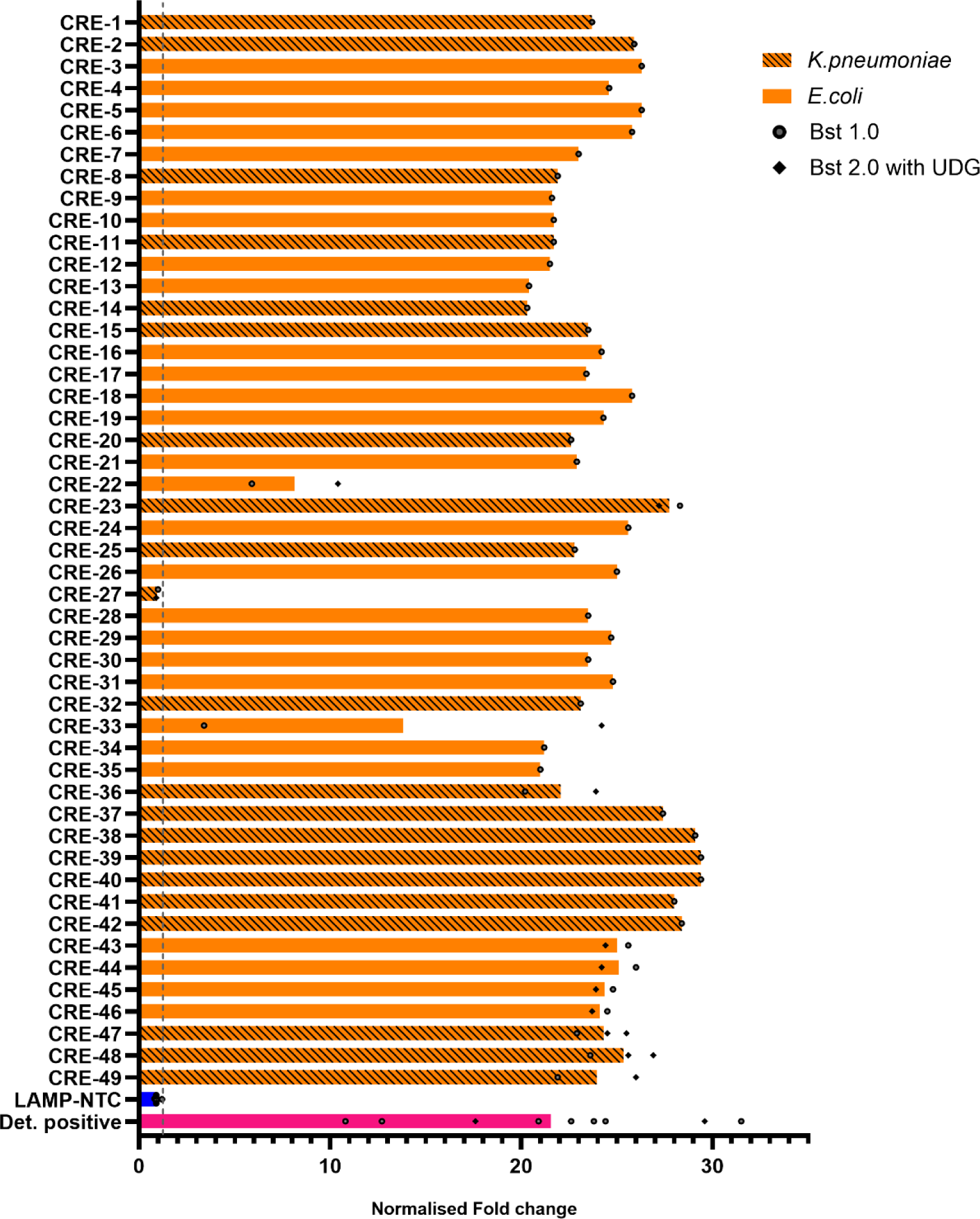
Tested *PathCrisp* assay on unknown CRE samples. 49 Carbapenem-resistant Enterobacteriaceae (CRE) isolates obtained from patient samples were tested for the presence of NDM using *PathCrisp* assay. The obtained RFU values are normalized to the detection negative of each batch and plotted here. For a few samples, the assay was repeated twice, once with Bst 1.0 (without UDG) depicted as circles in the graph and a second time with BST 2.0 (with UDG) depicted as diamonds in the graph. The average value of each sample in case of repetition is represented as a bar graph, while individual values are marked either as circles or diamonds. Grey dotted lines represent the cutoff to consider if the sample is positive, which is the average of RFU of LAMP-NTC plus 3 times Std. deviation. The 49 samples obtained were either *K.pneumoniae* (Striped bars) or *E.coli* (Solid coloured bar). In each batch, the negative control for LAMP is no-template control, depicted as LAMP-NTC. For detection controls, negative control, with no LAMP product, is shown as Det. negative in the graph. For positive control, pJET1.2_NDM was used, depicted as Det. positive.

The diagnostic testing accuracy of the *PathCrisp* assay and the PCR were evaluated against Sanger sequencing results which is considered the gold standard (Table 3). The sensitivity of the PCR test was 85.42% while the PathCrisp assay was 100%. The specificity of both tests was 100%.

**Table 3:** Diagnostic testing accuracy of *PathCrisp* assay: The *PathCrisp* assay of 49 CRE samples is compared with PCR-based detection for Sequence confirmed 49 CRE samples.

|  | Sensitivity<br>(in percentage) | Specificity<br>(in percentage) |
| --- | --- | --- |
| PCR based Gel<br>analysis | 85.42 | 100.00 |
| *PathCrisp*<br>assay | 100.00 | 100.00 |

### Testing *PathCrisp* assay on crude extract

In our study, the *PathCrisp* assay was validated using pure extracted DNA from clinical bacterial isolates, to reduce cost and further processes, we tested if the assay is compatible with crude extracts of DNA from the culture. For this, we transformed DH5⍺ *E. coli* cells with pJET1.2_NDM and used two types of samples (i) Colony from agar plate and (ii) overnight inoculated broth. In both cases, NDM-positive samples were successfully detected against negative controls, repeatedly (Fig 5).

**Figure 5:**
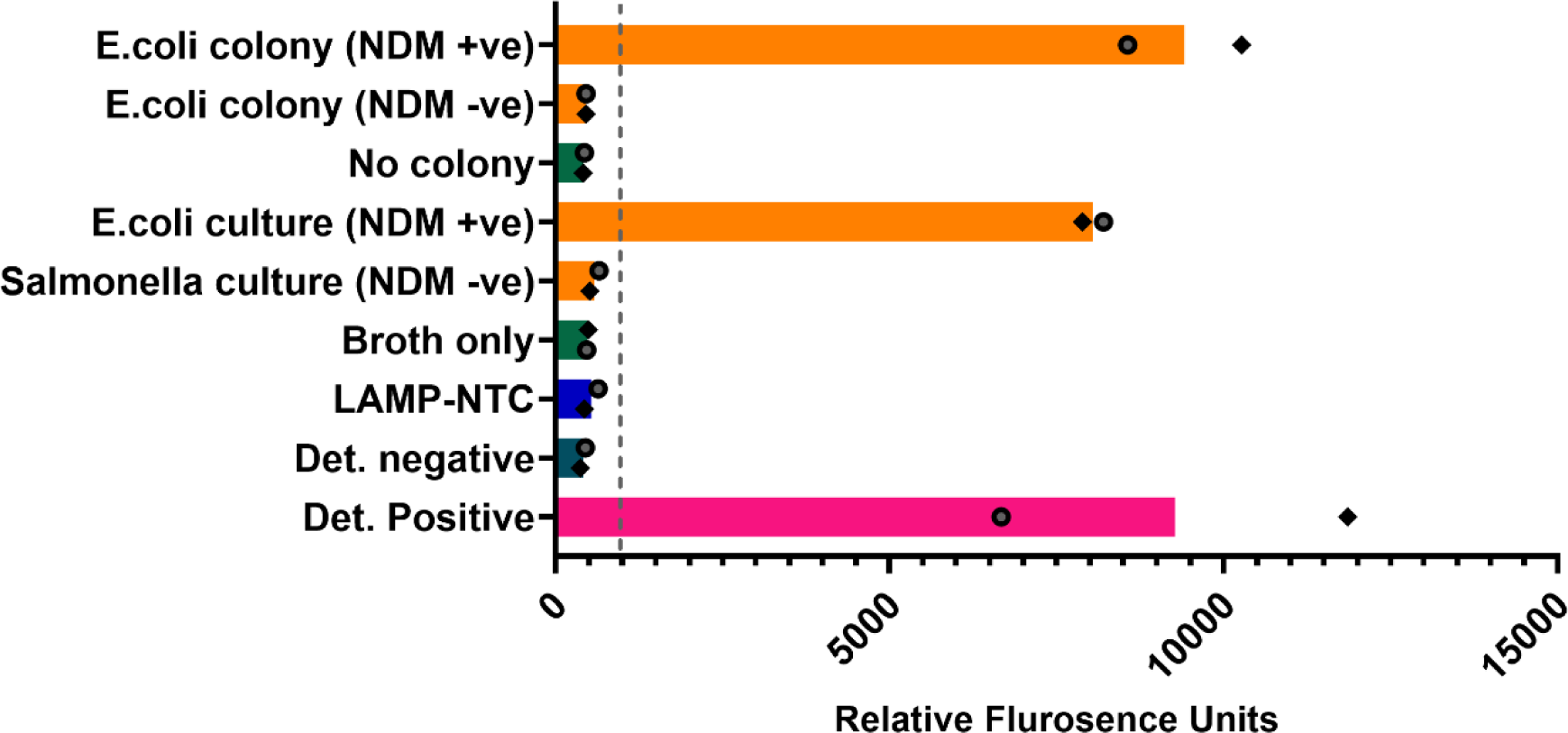
*PathCrisp* test on crude extract. pJET1.2_NDM transformed DH5⍺ *E. coli* cells were used to test the *PathCrisp* assay, after crude extraction. The assay was repeated twice, once with Bst1.0 (without UDG) depicted as circles in the graph and the second time with BST2.0 (with UDG) depicted as diamond shapes in the graph. The average value of each sample is represented as a bar graph while individual values are marked either as a circle or a diamond. The grey dotted lines represent the cutoff to consider if the sample is positive, which is the average of RFU of LAMP-NTC plus 3 times Standard deviation. Negative control for media was also included for each type of culture: LB Agar plate and LB broth, respectively. The negative control for LAMP is no-template control, depicted as LAMP-NTC. For detection controls, negative control, with no LAMP product, is shown as Det. negative in the graph. For positive control, 0.3nM pJET1.2_NDM was used, depicted as Det. positive.

## Discussion

Our *PathCrisp-*NDM assay, a Cas12a-based detection method, employs a carefully designed guide to target an NDM region conserved across variants. Notably, very recently Shin, Jiyong, et al (2024) published a PCR-amplified Cas12a-based detection method for NDM using a sgRNA similar to our *PathCrisp* guide^30^. Additionally, Curti, Lucia Ana et al. (2020), reported that the Cas12a-based NDM-gene detection capacity significantly increased with pre-amplified product compared to target DNA directly^31^. - Our *PathCrisp* assay includes LAMP-based amplification, known for its high sensitivity^12^. While direct LAMP-based methods for NDM detection have also been proposed to improve sensitivity^32,33^, using longer and multiple primers can compromise specificity. Therefore, we reported a combination, a LAMP-amplified CRISPR-Cas12a-based NDM diagnostics, supported by clinical sample testing, that benefits from isothermal amplification of LAMP and the precise secondary validation by CRISPR.

Previously, LAMP-based amplification followed by Cas12a-based detection has been proposed for NDM^34^. Here, the assay uses two different temperature conditions, LAMP at 65°C for amplification and trans-cleavage at 37°C for detection. Fuchs, Ryan T et al. (2022) compared different Cas12a enzymes and found both AsCas12a and LbCas12a, which we used, exhibit both cis and trans activity active at 48°C^35^. Furthermore, to improve the specificity of LAMP-based detection of *Neisseria meningitidis,* the LbCas12a-based Trans-cleavage assay was set up at 55°C by another group^36^. We advanced these studies by using a constant temperature of 60°C for both steps of our assay. This resulted in a sequential assay maintained at 60°C throughout, enabling the use of simple heating instruments like a water or heat block.

When the analytical sensitivity of different types of amplification methods was compared, LAMP was better than the gold standard PCR methods^12^. Additionally, the combination of Cas12a and LAMP is known for its high sensitivity and specificity^34^. Our *PathCrisp* assay was able to detect as low as 700 copies when tested in a serial dilution of target-carrying plasmid. For AMR markers present in plasmid form, it is challenging to determine the copy number per CFU of bacteria and accurately determine the Limit of detection. Often, these AMR genes exist in multiple copies per plasmid, and each plasmid can be present in multiple copies per bacterium. Given that, the current limit of 700 copies detected using a surrogate plasmid through our *PathCrisp* assay is indeed promising.

The PCR method, a gold standard molecular method, was used to screen clinical samples for NDM positives. The previously reported PCR method^26^, when analysed on gel electrophoresis, showed that some of the samples had either no band or a non-specific band. However, many of these discrepant samples were positive for NDM when the PCR was repeated with different sets of primers. Meanwhile, our *PathCrisp* assay outperformed the gold standard PCR method in sensitivity.

Most of the LAMP-CRISPR/CAS12a-based diagnostics proposed used kit-based DNA extraction for target detection ^37–39^. Many others have also proposed methods to extract DNA and increase the concentration of the target gene, such as immunocapture magnetic beads^40^ or directly boiling the culture^34^. In the present study, the latter method was used to prepare extracts, demonstrating that both types of culture: from plate and broth samples could be used as the template for the assay. This implies that for rapid diagnosis of NDM, patient samples could be cultured in relevant bacterial media and tested within a few hours of incubation. This could lead to a significant reduction in the time required to grow culture and eliminate the need for morphological analysis of colonies formed after several days of culture, which unfortunately continues to be the current dominant form of diagnosis.

In summary, we have developed a LAMP-CRISPR/Cas12a-based isothermal assay, *PathCrisp* for detecting the most notorious carbapenemase in South Asian countries, NDM. It outperformed the PCR method in detection and can detect low copy numbers. Furthermore, pretreated culture can be directly used for this analysis The *PathCrisp* assay was developed as a rapid, point-of-care test and is instrument-light and easy to set up. Although in the present study, fluorescence-based validation was used as a primary read-out, we are developing a lateral flow test for easy identification. Additionally, we will continue to extend this to the detection of other major carbapenemases and hope to build a robust diagnostic test to help clinicians decide on antibiotic therapy soon.

## Data Availability

All data produced in the present work are contained in the manuscript.

## Acknowledgements

We acknowledge Rockefeller Foundation Grant Number 2021 HTH 018 and financial aid by Axis Bank for funding the multi-centre AMR mapping and genomic analysis study at Ashoka University. Technical teams from Max Healthcare and Sahyadri Hospital for maintenance of pathogen culture stocks and DNA extractions. Yuvraj J. and Vasundhara K. for building the genomic assemblies. UGC-SAP-DRS-III, DST-FIST and DBT-BIF, Govt. of India for the infrastructural support to the Department of Biosciences, SSSIHL, Prasanthi Nilayam and ICMR-SRF from Govt. of India. Dr. BEP received extramural Research support from ICMR, Govt. of India. Abbireddy Sairam and Harinath Bapat for their support with clinical sample extraction used for assay validation. CRISPRBITS team for their unwavering support, and a special mention to Bhargav CN, Salil Hangekar and Kanikah Mehndiratta for their scientific inputs, constant troubleshooting help and helpful discussions. We would also like to acknowledge ex-team member Bilal Shah for the initiation of this project and Santosh M and Lavanya C for administrative support. Additionally, we also would like to acknowledge the Sequencing facility of the National Center for Biological Sciences and Barcode Biosciences for sequencing DNA samples. RA would like to thank Z, Z and K for their love, strength and support.

## Author Contribution

S.P., A.S., R.A., and V.G. contributed to the conceptual design of the study. S.P. conducted the development and validation of the PathCrisp assay with support from D.M., A.S., M.N., K.B.S. and V.H.. N.G., S.K., M.K., and B.T. sequenced and analyzed Indian clinical samples for designed oligos. J.R. and B.E.P provided clinical samples and extracted DNA for validating the PathCrisp. S.P., N.G., S.K. and R.A. performed the analyses and generated figures and tables. S.P. and R.A. wrote the manuscript. V.G., S.A. and V.C. provided input on subsequent drafts. All authors acknowledge full responsibility for the analyses and interpretation of the report.

